# The Impact on Social Well-Being of Cognitive Bias about Infectious Diseases

**DOI:** 10.1101/2024.02.25.24303338

**Authors:** Radomir Pestow

**Affiliations:** Mathematical Institute, University of Koblenz

## Abstract

We investigate the relationship between bias about the severity of infectious disease and social well-being.

First, we establish empirically the existence of bias and some of its causes; specifically, during the COVID-19 pandemic.

Second, we derive an integrated economic-epidemiological differential equation model from an agent-based model that combines myopic rational choice with infectious disease dynamics. Third, we characterize axiomatically a model of an ethical, impartial, eudaemonistic and individualist observer. We proof that such an observer evaluates the state of society (social welfare or well-being) according to the utilitarian principle.

Fourth, we show numerically, that while increased risk-perception indeed improves epidemiological outcomes such as peak of infections and total incidence, the impact on social well-being is ambiguous. This result urges to look beyond cases and deaths.

Finally, we point out three possible future research directions and highlight some critical issues that arise in the normative direction.

## 1 Introduction

In this paper we develop an economic-epidemiological model that can been seen as an answer to some of the challenges raised by (Dangerfield et al., 2022). As is stressed in (Dangerfield et al., 2022) the COVID-19 pandemic showed that a pandemic and the societal response to it can have far wider reaching consequences than only cases and deaths. The integration of epidemiological models with economic models designed to capture these consequences can therefore improve our understanding of the impact of epidemics on different aspects of society.

Two of the challenges discussed are first, the normative question of “how to capture the range of impacts of an intervention when evaluating policy?” (Dangerfield et al., 2022, challenge 1); and second, how to model the interaction between health risks and behavior (Dangerfield et al., 2022, challenge 2), specifically behavioral departures from norms of rationality?

The first question is addressed by building a new normative, axiomatic model of an ethical observer and thereby integrate social welfare analysis with epidemiology based on first principles (d’Aspermont in Arrow et al., 2002). As a result, the ethical judgment of the observer depends on the sum or, equivalently, average of well-being (which is precisely the utilitarian principle). This average at the same time quantifies the meaning of social/collective well-being or synonymously (social) welfare. Welfare analysis has already been used by (Aadland et al., 2021), but in a forward-looking way. In contrast to this the utilitarian principle can be applied retrospectively and aggregates the realized moment-to-moment experience of well-being of a (possibly heterogenous) population of hedonic agents.

The paper contributes to the second challenge by extending the SIR model (Kermack/McKendrick, 1927, or Martcheva, 2010 for a modern exposition) with agents that behave myopically rationally in the von Neumann-Morgenstern sense von Neumann, 1947. That is, agents have a sense of their own well-being, observe the negative health impacts of the contagious disease in their social environment, the risk of infection, their subjective avoidance costs and act accordingly as to maximize their well-being in a myopic way, i.e. only the present expected utility is maximized. This allows us to explore the effects of cognitive distortions, or biases, by which we mean the distortion of cognitive representations compared to objective reality, which is essentially the influential definition by (Haselton et al., 2015).

By (subjective) well-being we mean here happiness in an affective hedonic sense as in (cf. Y.-K. Ng, 2022). Cardinal measurability of well-being extended in time as required by our modeling is implied (at least in principle) by a) *intra*-personal comparability of levels and of differences in well-being, b) *inter*-personal comparability of well-being units (e.g. a just noticeable difference in well-being), and c) inter-temporal comparability of well-being in time (cf. Y.-K. Ng, 2022 Chapter 6.1 and references therein, cf.Kahneman, 1997)). Empirical well-being indicators on the other hand can be obtained by sampling moment to moment from experiences, monitoring of reward centers or stress hormones, empathic evaluation, willingness to pay, and in aggregated form from quality of life indicators, income, among others (cf. Y.-K. Ng, 2000, Y.-K. Ng, 2022, Chapter 6 and references therein, Kahneman/Krueger, 2006).

Finally, in addition to addressing modeling challenges, we explore the impact of bias on various epidemiological and, in particular, (social) welfare outcomes, at least in a theoretical setting. The biases we consider are driven by large extent by distorted media presentations of the severity of disease, as described in the observations below. In terms of natural selection, this sort of bias can be thought of as an artifact of our cognitive mechanisms being applied naively to the *evolutionary novel information environment* of our present time (cf. Haselton et al., 2015).

The impact on social well-being or welfare is relevant as bias inducing risk-communication was employed on purpose by some governments during the pandemic (see observations below), while at the same time, supposedly being committed to the common good (e.g. the German cabinet is required to swear an oath to devote themselves to well-being of the German people). The common good to which modern democracies are committed can be seen as an aggregate of individual well-being. One possible quantification of this is, as we will see, the sum of well-being.

Integrated behavior-disease models in mixed populations are briefly reviewed in (Wang Z. et al., 2015), and more recently in (Bedson et al., 2021). Other mixed population models with avoidance effort (in form of contact reductions or social distancing) and (forward looking) rational choice are treated in e.g. (Toxvaerd, 2022, Bhattacharyya/Reluga, 2019, Fenichel et al., 2022) among others. A rules-based approach that takes perceptions into account is presented in (Dönges et al., 2022). The myopic rationality approach we used here simplifies the model compared to the expected discounted utility approach but is still more grounded in theory than simple, ad-hoc, rules-based behavior. As such it can be seen as a compromise between the two approaches in terms of feasibility and plausibility.

Also related are models that take fear into account (Mandal et al., 2020, Juga et al., 2021, Mpeshe/Nyerere, 2021, Epstein et al., 2021, Jain et al., 2022, Retzlaff et al., 2022). However, fear understood as an emotion is not explicitly treated in our model. One may identify fear with the expectation of a decrease in well being from infection, but we will not make further use of that. For our purposes it is enough to focus on the relationship between perception and behavior without other intervening psychological variables.

A helpful pedagogical example in designing the model was (Tuckwell/Williams, 2007). The outline of the paper is as follows, in section 2.1 we will first state some motivating observations for the modeling, where we show the existence of bias and its (partial) causes. In section 2.2 we introduce our modeling assumptions for the epidemiological, behavioral and normative sub-models. The results are derived in section 3, which includes derivation of the mean field equations, the derivation of the utilitarian principle from the normative model, a partial qualitative analysis of the course of the pandemic, and finally the simulation results, where we scan part of the parameters space to evaluate the impact on social well-being of cognitive bias. We discuss the results in section 3.3.2 and conclude in section 5.

## 2 Materials and Methods

### 2.1 Observations

#### 2.1.1 Existence of Bias

Before asking what the consequences of bias are for well-being, we should first ask whether there exists any bias about infectious diseases, especially during the COVID-19 pandemic. In general, the very fact that there is political polarization on issues of fact (as opposed to issues of value) is evidence of the existence and persistence of bias among large groups of the population in different polities. For mutually exclusive positions cannot all be true at the same time; at least one of them must be false. It is likely that most readers, whatever their political persuasion, consider some of their political opponents (in private or public life) to be (heavily) biased or misinformed about the particular issue on which there is disagreement; and the feeling is just as likely to be mutual. But let’s be more specific. Early on, the WHO reported a fatality rate of 3.4% (WHO, 2020) for COVID-19 and contrasted this with a fatality rate of “well below” 1% for seasonal influenza, claiming that only 1% of coronavirus infections are asymptomatic. However, subsequent seroprevalence studies found infection-fatality rates ranging from 0.00% to 1.63%, with a median of 0.27%, as, incidentally summarized in another WHO publication (Ioannidis, 2020a).^2^. A later reconciliation of six systematic evaluations of pre-vaccine seroprevalence studies settled on an infection fatality rate of around 0.15% (Ioannidis, 2021). In any case, this early announcement can be seen as a very prominent and influential example of bias^3^.

Given that there was already bias in the expert community, it is not surprising that there was also bias in the general population, as it is influenced by the expert community. For example, the well-executed COSMO study (Betsch et. al., 2020) shows a wide divergence of opinion in the population about how many are in the risk group, ranging from 0% to 100%, with pronounced local peaks at “round” decimal numbers such as 5, 10, 15, 20, 30, 40, 50, 60 (COSMO, 2022). A Gallup poll (Rothwell et al., 2021) found that 92% of US adults and 62% overestimated the risk of hospitalization for the unvaccinated and vaccinated, respectively. Another online questionnaire in late February 2020 found that “US and UK participants’ median estimate for the probability of a fatal disease course among those infected with severe acute respiratory syndrome coronavirus 2 (SARS-CoV-2) was 5.0% (IQR 2.0%-15.0%) and 3.0% (IQR 2.0%-10.0%), respectively.” (Geldsetzer, 2020).

#### 2.1.2 Causes of Bias about COVID-19

Having established the existence of bias towards infectious diseases, we will briefly consider some of its contributing causes.

##### Immediate Causes of Bias

Obviously, information beyond our immediate sense experience, our memory thereof, and our inferences therefrom must reach us through social channels, of which the organized media are a part. Since a particular virus is beyond immediate sense experience, the vast majority of people form their beliefs about viruses in general, and coronaviruses in particular, on the basis of socially transmitted ideas, many of them through organized media. It is therefore clear that these opinions are a reflection of the social milieu (real and virtual).

Now, a content analysis of the UK media coverage between January and May 2020 found that “journalists relied heavily on fear-inducing messages by emphasizing threats related to COVID-19 and, though to a lesser degree, measures against these threats” (Hase et al., 2022). Another content analysis of a global media sample came to a similar conclusion that “Human interest and fear/scaremongering frames dominated the global media coverage of the pandemic.” (Ogbodo et al., 2020). Finally, COSMO reported that around 40-50% feared the Coronavirus, and that 40-60% thought more or less often about the disease throughout 2020 (COSMO, 2022); which, as argued above, mostly reflects social milieu and, in light of the previous argument, the type and prevalence of media content. We will not go into further details as these findings of predominantly negative media messaging at the beginning of the pandemic are probably in line with the personal experiences of most of our contemporaries in Western societies.

As we said above, media constitutes only one part of the informational milieu of a given individual. Obviously, others parts of the social milieu have an influence on the formation of beliefs too. Also, the causality is not one-way (i.e. not only from media to beliefs), but individuals choose which media they consume and which other individuals the associate with (thus, beliefs imply choice of media and milieu in general).

##### Mediate Causes of Bias

Pursuing this causal thread leads us to the role of governments in the pandemic. There are indications that exaggerated risk-perceptions were actually intended^4 5^ and promoted^6^ by some governments in order to increase compliance with pandemic measures and thereby reduce the spread of infections. Some influential publishers in German speaking countries followed the government lead (with good intentions probably) ^7 8^ in order to spread the Government message.

Other prominent state of the art recommendations of behavioral change tactics aimed to increase socially induced conformism^9^ and mental rigidity^10^ (Bavel et al., 2020)^11^.

This artificially reinforced a narrow focus on mortality and morbidity of one disease and lead to a loss of focus on other societal problems (which include more than only diseases), as well as to a disregard of collateral damages (Schippers, 2020). The narrow focus on deaths and infections was also reflected in influential modeling that disregarded all consequences beyond infections and deaths (Joffe, 2021).

Here again we want to stress, that the causality from politics to media to beliefs is not only in one direction, but that it is rather a complex interaction between and within voters, media, politics and other institutions.

### 2.2 Model

#### 2.2.1 Epidemiological Assumptions

We consider a population of homogeneous agents, *N*, that is subdivided into three distinct classes: susceptibles *S*(*t*), infected *I*(*t*), and recovered *R*(*t*). At each time step, susceptibles choose the level of effort *a ∈* [0, 1] they will make in order to avoid an infection (measured in terms of a reduction in transmission probability). After that, each susceptible makes one random contact uniformly distributed over the whole population. If agents make contact with an infected individual, they get infected with probability *β*(1 *− a*). Infected agents recover with probability *γ*, and recovered agents stay recovered.

Since the agents are homogeneous, they all have the same information and react in the same way to this information. Therefore all susceptibles will choose the same action *a*.

What are the expected changes in the numbers of the compartments? Let *S*(*t*), *I*(*t*), *R*(*t*) and the action *a* be given, then

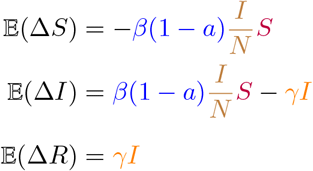

Here the fraction of infected 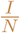 is equal to the probability that a susceptible agent will meet an infected (due to the uniform distribution of contacts). Further, *β*(1 *− a*) is the transmission probability, *β*, reduced by the avoidance effort. Each susceptible agent has therefore a chance of 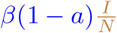 to get infected (getting infected is a Bernoulli variable). Therefore, in sum, 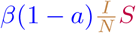 agents are expected to get infected on average.

Similarly, recovering is a Bernoulli random variable with probability *γ*. Therefore, as many as *γI* are expected to recover.

#### 2.2.2 Behavioral Assumptions

We additionally assume that the agents have a sense of their own well-being and act accordingly with myopic von-Neumann-Morgenstern rationality (von Neumann, 1947, Marschak, 1950, Harsyani, 1955). That is, the agents choose that action which *maximizes their present expected value* (in contrast to agents with a finite time horizon, that would maximize the expected, and possibly discounted, present value in their time horizon).

Effort to avoid an infection bears a cost *c*(*a*), which is zero if no effort is exerted, *c*(0) = 0, has constantly increasing marginal costs *c*″(*a*) = *κ >* 0 (for some constant *κ*), and is evaluated in terms of the agents well-being. We further assume that the first additional unit of protection has no additional cost in terms of well-being, *c*′(0) = 0. Getting an infection on the other hand bears a probabilistic health cost, with distribution Θ and mean 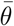, for the agents well-being.

The agents observe these health outcomes in the population, possibly with biased exposure or selective attention, so that some outcomes are over-/under-sampled, resulting in the actually observed distribution 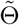 with the shifted perceived mean 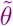.

The agents have the following information available: effort costs *c*, perceived health costs 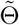, effective transmission probability *β*(1 *− a*) and the probability of meeting an infected 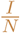. We also assume that the costs are additive.

Taken all of the above together, the agents thus try to choose the least worst expected outcome in the following decision problem

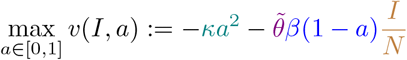

where *κa*^2^ are the protection costs and 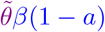 is the expected health cost for that protection level both evaluated in terms of well-being. Since the value function is homogeneous in the cost parameters, we can set *κ* = 1 without loss of generality (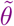 reads then as the health cost in proportion to the protection cost parameter). The optimal effort level is therefore 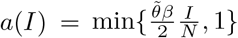.

The behavioral model conforms to observations that perceptions of increased risk about infectious disease promote risk-avoiding behaviors (e.g. Floyd et al., 2000, Majid et al., 2020, De Bruin/Bennet, 2020).

#### 2.2.3 Value Assumptions

For the purpose of making a value judgement on the state of the society of agents (i.e. a judgement of the form “societal state x is better than societal state y”), we will characterize how an ethical, impartial, eudaimonistic (in the original sense as in well-spirited), and individualist observer forms a judgement on the comparative value of two societal states (cf. d’Aspermont in Arrow et al., 2002, Y.-K. Ng, 2000). The following axioms (esp. Eudaimonism and Individualism) are formally related to those of pure utilitarianism as described in (d’Aspermont in Arrow et al., 2002). Here we choose utilitarianism over the other prominent alternative, namely Rawlsian or egalitarian social justice, simply for a formal reason: in an infinite population, that we are going to consider, the welfare level of the least-well off is always the same, i.e. a constant (the worst disease outcome is always realized in an infinite population). This trivializes the concept. Besides, the average well-being is already an interesting measure in itself.

Let then *x, y ∈ C* be social states in the set of comparable states *C*, and let *P* be the social ordering relation of the impartial observer, were *xPy* means that *x* is considered to be a strictly better social state than *y*. Analogously *I* is the indifference relation of the observer between two social states, where *I* and *P* are compatible with each other. Furthermore let *w*_*i*_(*x*) denote the well-being of agent *i* in social state *x* and *w*(*x*) the vector of well-being of all the agents. Then the observer is assumed to be guided by the following principles:

##### Unanimity (or Pareto Principle)

If *all* agents are better off in one social state than in another with one individual being at least strictly better off, then the former state is strictly preferred by the ethical observer: [if [*w*(*x*) ≥ *w*(*y*) (compared componentwise) and one *i* with *w*_*i*_(*x*) *> w*_*i*_(*y*)] then *xPy*].

##### Impartiality

All agents are treated equal. The well-being of one agent is not considered more important than that of others: [let *w*_*π*(*i*)_(*x*) = *w*_*i*_(*y*) with permutation, *π* then *xIy*].

If the observer *were* to prefer individual *i* to individual *j*, he would prefer a given state *x* over a given state *y*, if *w*_*i*_(*x*) = *a, w*_*j*_(*x*) = *b, w*_*i*_(*y*) = *b, w*_*j*_(*y*) = *a*, and *a > b*; i.e. he prefers the state were individual *i* gets the higher level of two possible levels of well-being rather than individual *j*.

##### Eudaimonism

The observer is indifferent between two states, if the individuals have the same well-being in both states, i.e.: if *w*(*x*) = *w*(*y*), then *xIy*.

Thus, the observer does not care about anything extraneous to well-being, such as the type or color of clothes one wears, but only insofar it affects well-being.^12^

##### Individualism

When comparing two social states, the judgement depends only on the difference in well-being between these two states: let *x*_1_, *x*_2_ and *y*_1_, *y*_2_ be social states with *w*(*x*_1_) *− w*(*y*_1_) = *w*(*x*_2_)*− w*(*y*_2_) then *u*(*x*_1_)*Pu*(*y*_1_) *⇔ u*(*x*_2_)*Pu*(*y*_2_) and *u*(*x*_1_)*Iu*(*y*_1_) *⇔ u*(*x*_2_)*Iu*(*y*_2_). In words, if the difference in one pair of states is the same as the difference in another pair of states, then the two pairs of states are treated the same.

Here, each individual is considered to be a world unto itself, as if the individuals were living on different planets, since relative levels of well-being are not taken into account. Note, however, that empathy (where the well-being of one agent somehow depends or is related to the well-being of others) and Weber-Fechner-like properties of well-being lead to more egalitarian conclusions even in this individualistic setting.

##### Maximum Domain

The set of social states *S* which the observer can compare includes all possible distributions of well-being, i.e. ℝ^*n*^ = *w*(*S*). This means that for every well-being distribution one can think of, there is a social state that realizes it.

The social welfare ordering of the impartial observer is then given by Σ_*i*_ *w*_*i*_(*x*) in the following sense (see the appendix for the proof)^13^:

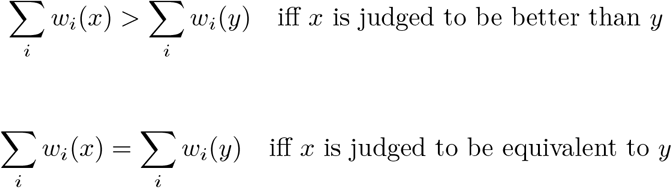

Note also that the strict preference relation defined by *P* := {(*x, y*) *∈ S*|Σ_*i*_ *w*_*i*_(*x*) *>* Σ_*i*_ *w*_*i*_(*y*)}, and the indifference relation defined by *I* := {(*x, y*) *∈ S*| Σ_*i*_ *w*_*i*_(*x*) = Σ_*i*_ *w*_*i*_(*y*)}, both fulfill the axioms, thus proving the consistency of the axiom set.^14^

In our case we consider the total well-being *w*_*i*_ of an agent *i* to be the episodic or cumulative reward, i.e. 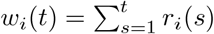 (cf. Kahneman, 1997

The expected change in per-capita welfare, 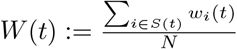, is thus given by

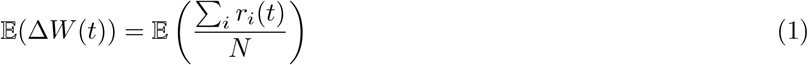

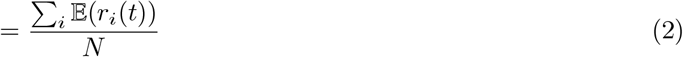

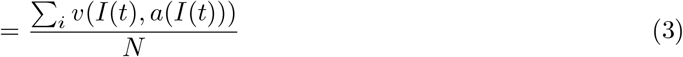

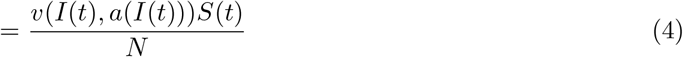

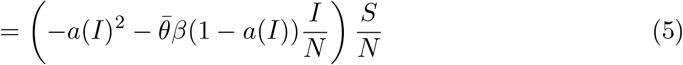

where the index *i* ranges over the susceptible agents.

Here, *v* is the expected value of action *a*, given the number of infected *I*. In the last line, *−a*(*I*)^2^ is the effort cost (which is the same for all susceptible agents since all take the same action). Further, a 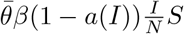 is the aggregated true health cost, as 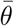 is the true mean of the health costs.

## 3 Results

### 3.1 Limit Model

If we normalize the population, and assume that the time steps were sufficiently small, we obtain the following differential equation system in the infinite population limit by the Law of Large Numbers:

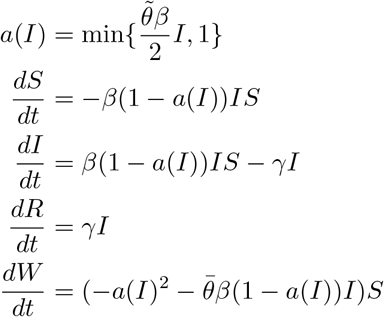

where 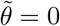 gives us the standard SIR-Model.

Some observations are in order. First note, if *I*(0) is such that 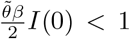, then 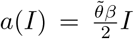 throughout the whole pandemic, which simplifies the model further. This is the case, because *a*(*I*) cannot grow above 1. As *a*(*I*) is continuous (in *I* and therefore in *t*) it would need to be equal to 1 at some point, if it were to grow above one. But then *β*(1 *− a*(*I*))*IS* would be equal to 0 in the first and second equation and the number of infected would fall, pushing *a*(*I*) under 1 again. The system will then have the following form for appropriate starting values

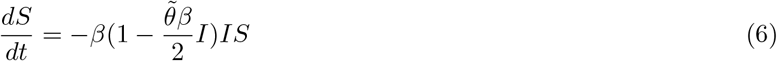

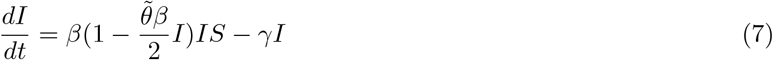

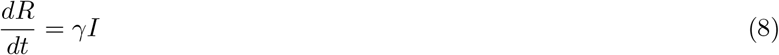

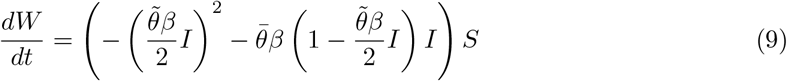

Another special case is where 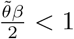, then we will also have *a*(*I*) *<* 1 through out and 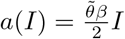

### 3.2 Qualitative Analysis

We see that if *I*(0) ≈ 0, then *a*(*I*) ≈ 0, and the dynamics are nearly the same as in the basic SIR-model. Thus, we can say that right at the start, when the number of infections is small, both models give nearly the same results.

Further, *S*′ is negative throughout. Therefore, *S* is monotonically decreasing and bounded from below (since *S* is stationary in *S* = 0 which implies that *S*′ = 0 at that point, i.e. it will stay there and cannot cross it). Thus *S* has a lower limit.

Now assume that *I*(0) is small at the start and consider the growth rates of *S* and *I* (with 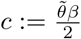):

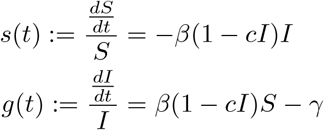

Here we note that *g* can cross 0 only from above but not from below. For assume, *g*(*t*_0_) = 0, then *I*′ = 0. Now, if we take the derivative of *g*, we see that *g*′ = *−βcI*′*S* +*β*(1*−cI*)*S*′ = *β*(1*−cI*)*S*′ *<* 0. Therefore once *g* is below zero, it will stay below zero and the number of infected will be continuously falling.

Again, if *g* is positive, then *I*′ *>* 0. Taking the derivative, we see that *g*′ will be negative, since both *−βcI*′*S* and *β*(1 *− cI*)*S*′ will negative. Therefore, the growth rate will be continuously falling and correspondingly the growth of *I* will be continuously slowing down.

We get the following picture. If *I*(0) is small enough, the model will behave in the beginning as the SIR model. The growth of *I* will continuously level off until *I* reaches, presumably, its peak, after which *I* will be continuously falling, and *I* and *S* will level off to their limits.

### 3.3 Simulation Results

To get a more clearer picture, we will take a look at some simulation results; first at the epidemiological outcomes, then at the welfare outcomes.^15^

#### 3.3.1 Epidemiological Outcomes

Note, that the epidemiological outcomes depend only on the perception of the disease, while welfare depends on both the perception and the actual severity. For the epidemiological outcomes we will therefore take a look at how disease perception, 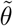, affects the course of the epidemic, especially the peak of infections, *I*_*m*_ := max_*t*_ *I*(*t*), time of peak, *t*_*m*_ := arg max_*t*_ *I*(*t*), total incidence, *f* := lim_*t→∞*_ *R*(*t*), and duration, *d* := inf{*t*|*I*(*t*) *< I*(0)} (i.e. the first time, when the number of infected is below the initial value).

The figure 1 depicts how the epidemic as a whole is affected by disease perception in the short term.

**Figure 1:**
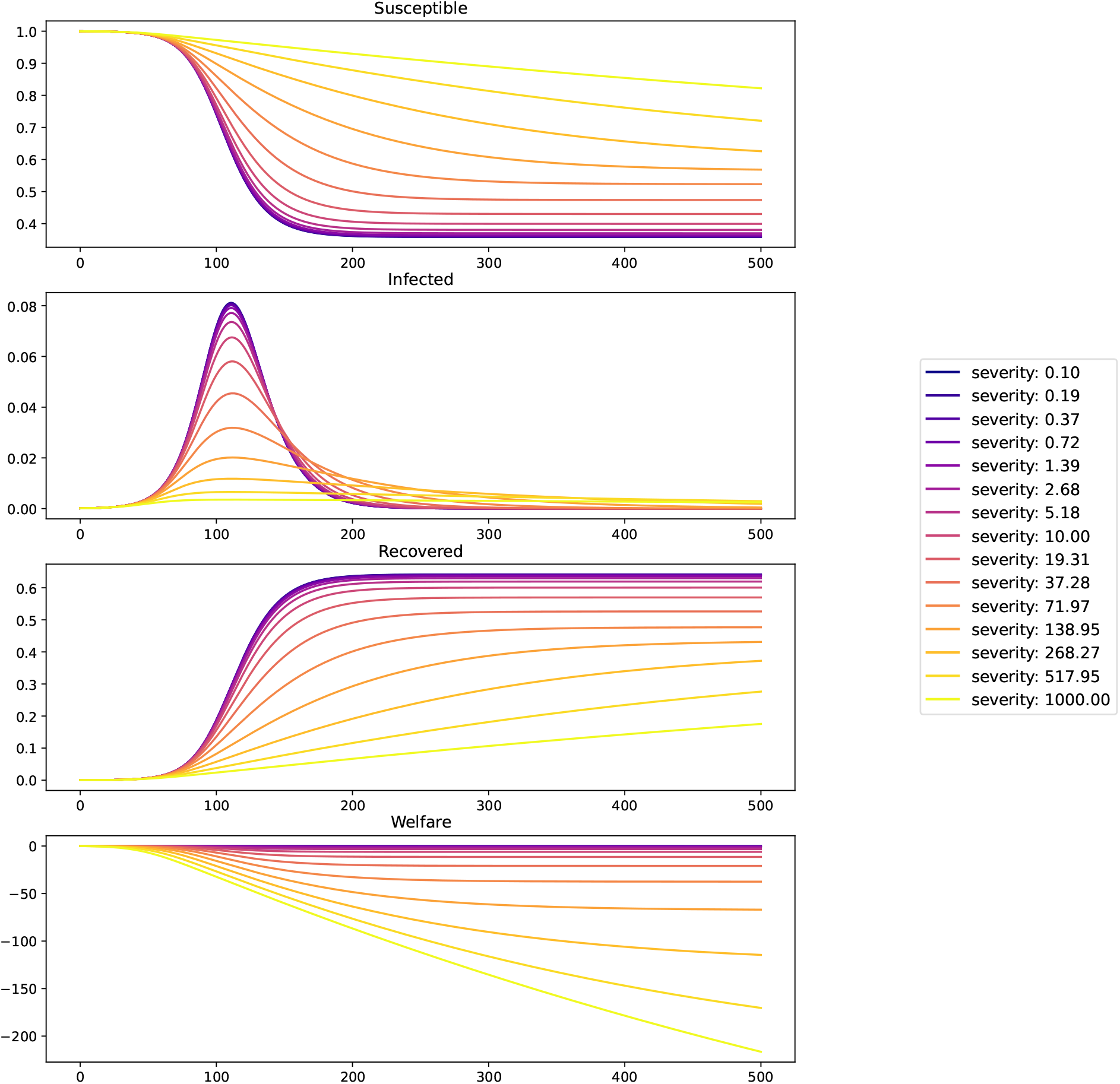
The epidemic as it develops in the short term with *β* = 0.2, *γ* = 0.125, *b* = 1, *I*(0) = 0.0001

These figures suggest that the peak of infections is increasing with perceived disease severity, while the final incidence as well as welfare are decreasing in severeness of the disease, while at the same time the duration is increasing. We will now take a look at the long term outcomes.

Here we took a long enough time frame (here *T* = 5000) so that the epidemics with the differing severity finish (in the sense that *I*(*T*) *< I*(0)). Final incidence is then simply, *R*(*T*).

Figure 2 shows us the peak of infections and the time of their occurring. Figure 3 depicts the final incidence and duration of the epidemic.

**Figure 2:**
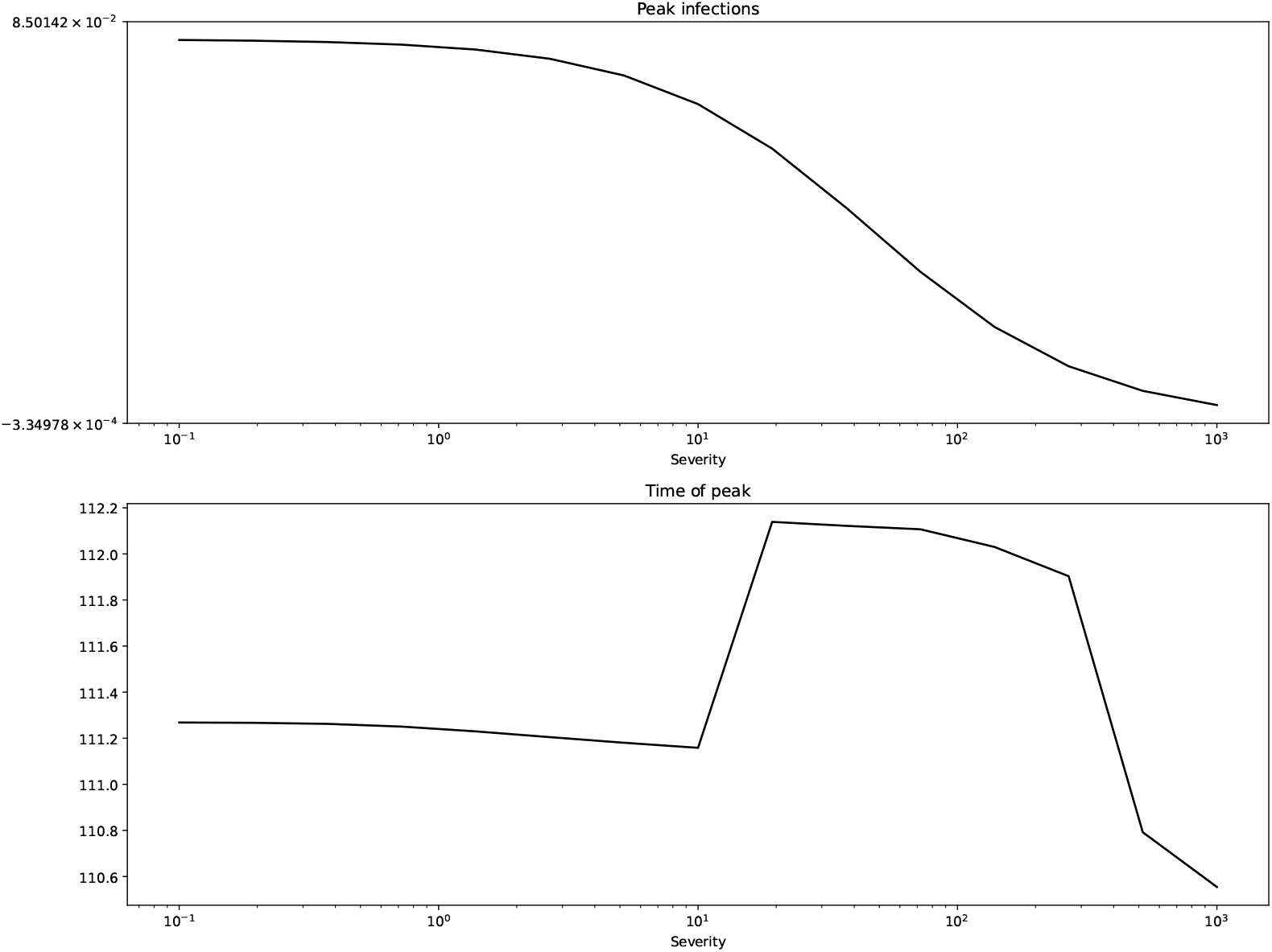
Peak of infections and time of peak for *β* = 0.2, *γ* = 0.125, *b* = 1, *I*(0) = 0.0001

**Figure 3:**
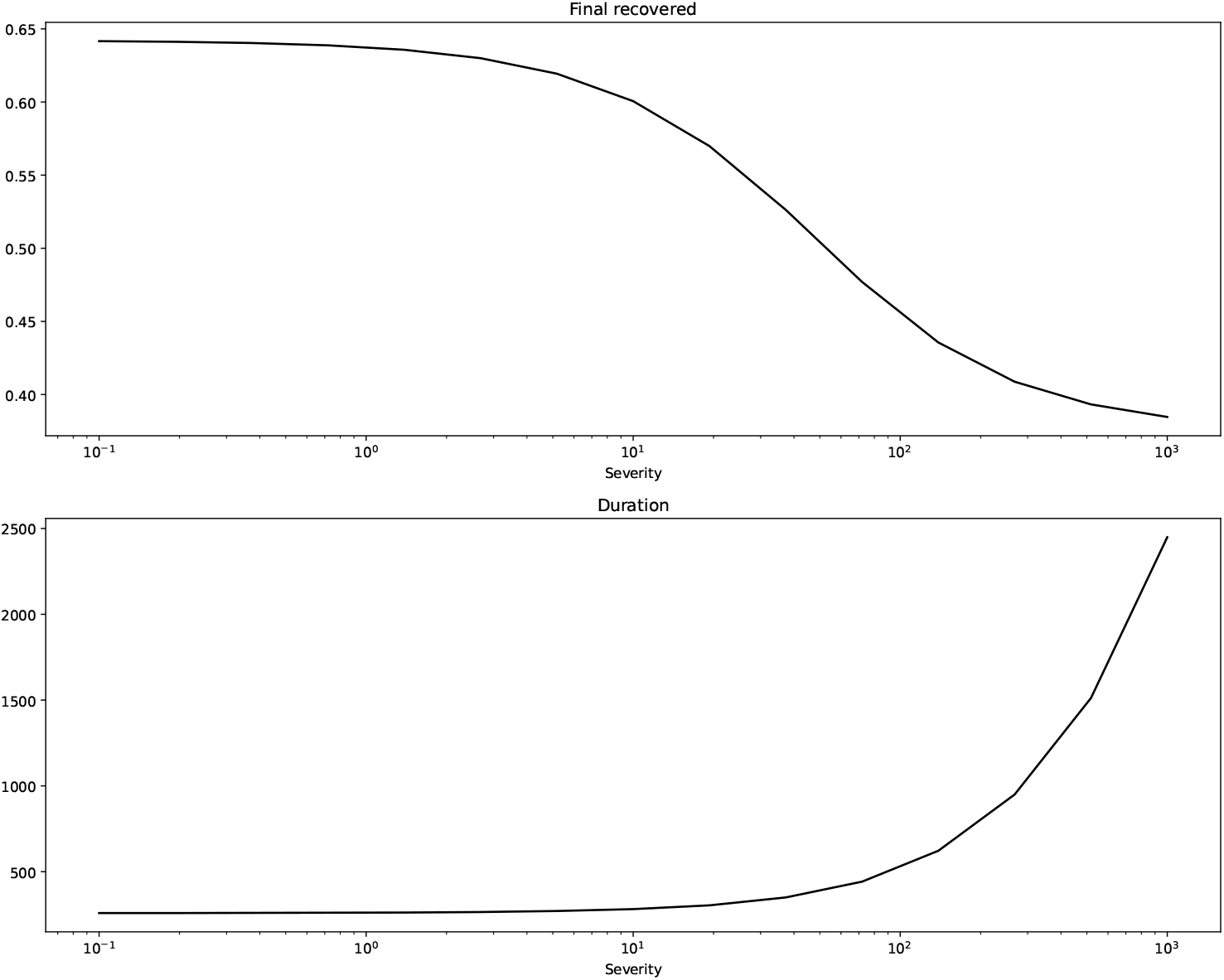
Final incidence and duration for *β* = 0.2, *γ* = 0.125, *b* = 1, *I*(0) = 0.0001

The figures confirm what we suspected from the short term depictions: peak of infections and final incidence are monotonically decreasing in severity while the duration is monotonically increasing.^16^

Does monotonicity hold for all parameters values? To answer this, we evaluate the model on a parameter grid of *N*·*M* points with 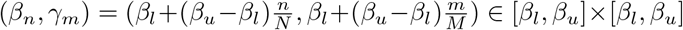 On each grid point we sampled perceived severity on *P* points with 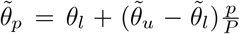 and checked the outcomes, 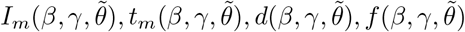 for monotonicity and convexity in 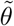. Here 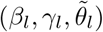 are the lower bounds and 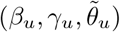 are the upper bounds, which we take to be (0.1, 0.1, 0) and (1, 0.9, 100) respectively with *N* = *M* = 10 and *P* = 20 Figure 4 show the monotonicity and convexity regions in epidemiological parameter space for the various outcomes Ω *∈* {*I*_*m*_, *t*_*m*_, *d, f*} with the following color coding:

**Figure 4:**
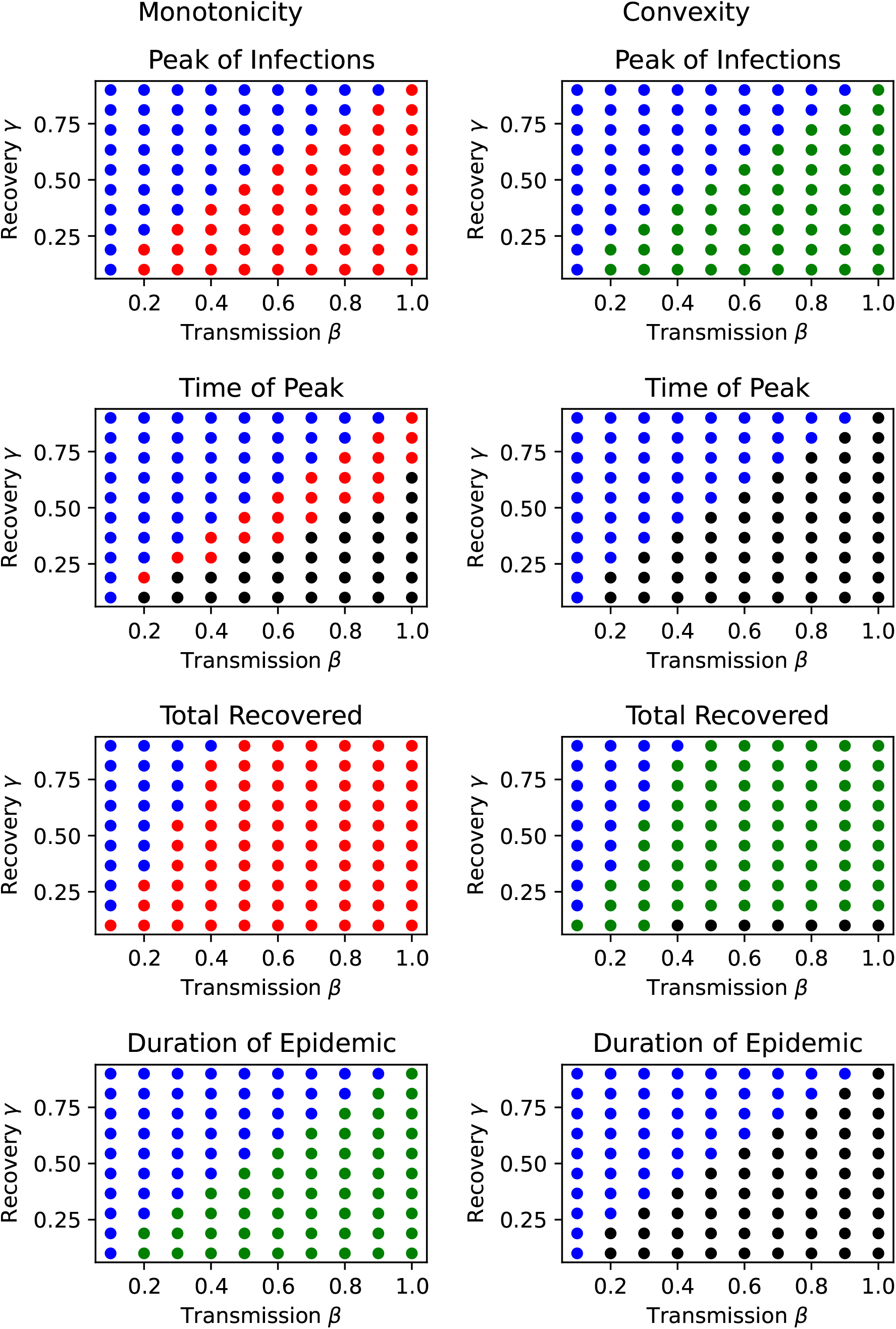
Monotonicity and Convexity results

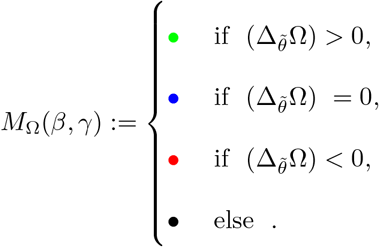

where the inequalities mean that the differences have to be strictly greater than zero for at least one value of 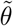. For the numerical evaluation we first checked whether the differences are nearly zero and then checked for the inequalities.

Here we see that the effect of threat perception is unequivocal with respect to peak of infections and final incidence (both constant or monotonically decreasing with increasing threat perception), while the duration of the pandemic is either (nearly) linear or increasing with threat perception. In addition the peak of infections are (nearly) linear or convex in threat perception.

#### 3.3.2 Welfare Outcomes

For the simulation, we first solved the epidemiological model for a given level of perceived disease severity and then integrated the welfare equation with the various real disease severities. The real severity ranged here from 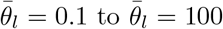 on *Q* points with 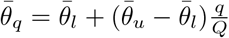. From that we calculated similar to above how final welfare *W*_*∞*_ := lim_*t→∞*_ *W* (*t*) is affected by varying the perceived threat levels for a given real threat level. Thus *N* · *M* · *Q* points were sampled and the first and second differences in Welfare (as above) were evaluated by varying the perceived threat levels. Table 1 depicts the aggregated results.

**Table 1:** Percentages of the monotonicity and convexity regions in the parameter region for *β*, *γ*, 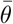.

| | $> 0$ | $= 0$ | $< 0$ | other |
| --- | --- | --- | --- | --- |
| $\Delta W_\infty$ | 30.3% | 0.0% | 21.5% | 48.2% |
| $\Delta^2 W_\infty$ | 21.5% | 30.3% | 0.0% | 48.2% |

We see that the effects of bias on final welfare are ambiguous. There are scenarios, where exaggerated risk-perceptions are welfare-increasing (first row, first column), and where it is the other way round (first row, last column). In other words, social welfare is not monotonically increasing in threat perception per se.

## 4 Discussion

In terms of the challenges mentioned in the introduction, the social welfare concept is certainly able to capture all relevant impacts as long as they can be related to the one common measure of well-being. The concept, as presented here, may at first seem simple, plausible and attractive. Nonetheless, utilitarianism has some issues of its own. Among those are so called “utility monsters”, i.e. “who get enormously greater gains in utility from any sacrifice of others than these others lose. For, unacceptably, the theory seems to require that we all be sacrificed in the monster’s maw, in order to increase total utility.” (Nozick, 1974, p.41). Thus, the theory certainly needs more refinement, and must be to applied judiciously. At least, there are no utility monsters in a homogeneous population such as in our model, although it is still a problem in principle.^17^ The second challenge, on the other hand, about behavioral departures from norms of rationality is easily accommodated within the von Neumann-Morgenstern framework, at least for distorted perceptions.

The rational choice and social welfare approach presented here are very flexible and allow to extend the modeling in many directions (see also below) as the vastness of the economic literature building on these concepts shows. For example, it is possible to add various social dynamics (opinion dynamics, political mechanisms) to the epidemiological model.

Regarding the effects of bias, our model confirms that increased risk-perceptions can indeed improve epidemiological measures like peak of infections and total incidence. However, increased risk-perception is not conducive to improvements in social welfare per se. This is, an improvement of epidemiological outcomes does not necessarily lead to an increase in social welfare or the average well-being in the population. That means that one cannot unconditionally recommend measures that increase risk perception, since prolonged, protective effort can create a burden that is larger than the burden of the disease itself. Rather, it necessary to take (social and individual) well-being holistically into account and it is not enough to focus only on epidemiological outcomes alone, which emphasizes the point made by (Dangerfield et al., 2022). One may be led to think that this is already clear by itself, but apparently it was not clear enough during the pandemic.

Additionally, in disease prevention, the costs of protection must always be weighed against the health benefits, which in turn requires timely data on perceived and real threat levels, as well as costs and benefits, preferably in terms of well-being; supposing that the controlling agency aims improve the welfare of the population.

But what the model also shows is that even unbiased, myopically rational agents behave in a way that is not always optimal from a social welfare perspective as there are scenarios where a deviation from unbiased perception improves welfare-outcomes (Table 1, first row of the first and third column).

## 5 Conclusions

This last point naturally raises the question of how public perception *should* be “optimally controlled” (in a mathematical sense), or influenced, in order to serve (social) well-being, or more general, the common good.

However, we feel that it is important to state some of the fundamental issues that are raised when addressing this question as they restrict the (ethical) applicability of optimal control to public perception.

First, it questionable whether the “state-manufactured consent of the governed” can bestow democratic legitimacy (in a subjective and objective sense) on those who govern. For it sounds absurd to say that a population that is psychologically ruled by the government is actually ruling the government. It rather seems, that the more a population is ruled by a government, the less it is actually a democracy, because psychological control by the government shifts the objective locus of control from the population to those in power (cf. Thomson/Ip, 2000, Christian/Bajaj, 2022). Second, this perceived loss of democratic control may lead to resentment and defiance, thereby contributing to political polarization, which in turn can become problematic when the threat is real and severe. Whether perceptions like this actually contributed resentment and thereby to polarization during the COVID-19 pandemic may well be an open question for empirical research. Third, a “doom loop” (to borrow a phrase by Laura Dodsworth, Rayner (2021)) can lead to persistently, sub-optimal and overly risky decision making procedures on the individual and societal level (due to panic decision making).

Fourth, if a government has the power to psychologically influence the population (as the Covid-19 crisis demonstrated that it has), there is a potential risk of abuse, that must be addressed.^18^ Fifth, closing the minds of the population to other opinions or steering them to specific opinions presupposes that one is in possession of the true opinion. This is, however, in a strict philosophical sense impossible pertaining to matters empirical. For, as Descartes argued, we cannot distinguish, as seen from the interior perspective, whether we are perceiving a realistic dream or actual reality. Thus, to believe in our sense experience requires already a leap of faith. To believe in the experiences of others requires many more leaps of faith. Philosophically speaking, all empirical science is doubtful; or in other words: empirical science cannot reach absolute certainty on the level of Mathematics.^19^

But more practically speaking, there is always the possibility of error. As we have shown at the beginning of the article, there are factual disagreements between large parts of the population, which proves that large parts of the population can be in error. There is no reason why the majority or the ruling class should be immune to this.^20^

Although the ethical and epistemic issues mentioned above may be tangential to our main research question, they are very important and essential to keep in mind if one chooses to pursue the research in a normative direction. That is, the above issues must somehow be reflected when deriving policy recommendations from the optimal control question. There are, of course, ethical ways of influencing the public, but creating misperceptions is certainly not one of them.

Another possibility is to take the theoretical research along a more descriptive route and extend the model to include various interacting social groups and institutions. The descriptive question, corresponding to the normative question above, would be: how do public perception and political decision-making *actually* interact, and what is the impact of this interaction on (social) well-being? Lastly, on the empirical side on might ask how to operationalize the variables of the model so as to make it predictive and testable. Conceptually, the model predicts that increasing risk-perception decreases the peak of infections and the total incidence, but increases the duration of the epidemic. These relationships seem very plausible, but is it possible to somehow test them? If they were not to hold, what other factors are intervening and how can they be accommodated within the model?

## A Proof of the Utilitarian Principle

We now want to proof from the axioms that

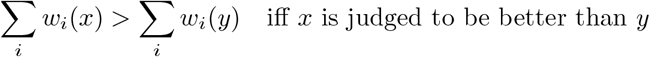

and

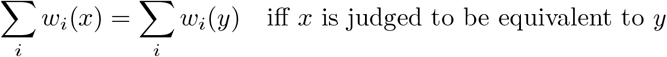

The proof for that borrows ideas from (d’Aspermont in Arrow et al., 2002).

We will first show the “ ⟹ “-part for both statements as this also implies the other direction. Let *J ∈* {*I, P*} denote the comparative judgment of the observer.

### 1. The following construction *C*

*S*×*S* ↦ *S*×*S* will create another pair of states, *u, v* so that *uJv* for given *x, y* with *xJ y*, i.e.: if C(x,y) = (u,v), and *xJ y*, then *uJv*, for *J ∈* {*I, P*}. Furthermore, if Σ_*i*_ *w*_*i*_(*x*) = Σ_*i*_ *w*_*i*_(*y*), then Σ_*i*_ *w*_*i*_(*u*) = Σ_*i*_ *w*_*i*_(*v*), i.e. *C* preserves the comparative judgement of the observer. Finally, the total number of zero-components in both *w*(*u*), *w*(*v*) is larger than the number of zero-components in *w*(*x*), *w*(*y*).

#### 1.2 Construction

Let xJy. First, rearrange the components of *w*(*x*), *w*(*y*) in decreasing order resulting in well-being vectors *w*′_*x*_, *w*′_*y*_, with permutations *π*_*x*_, *π*_*y*_. By (Maximum Domain) we can choose two states *x*′, *y*′ so that *w*(*x*′) = *w*′_*x*_, and *w*(*y*′) = *w*′_*y*_.

By (Impartiality), we have *xIx*′ and *yIy*′. Therefore by the compatability of *P* and *I*, the observer makes the same judgment on *x*′, *y*′ as on *x, y*, i.e. *x*′*Jy*′.

Now construct a new vector *s*, so that *s*_*i*_ is equal to the smaller value of *w*_*i*_(*x*′), *w*_*i*_(*y*′). Choose with (Maximum Domain) two new states *u, v*, so that *w*(*u*) = *w*(*x*′) *− s* and *w*(*v*) = *w*(*y*′) *− s*.

#### 1.3 Observation

By (Individualism) we have *uJv*, as the difference in well-being between *u, v* is the same as in between *x*′, *y*′, i.e. *w*(*u*) *− w*(*v*) = *w*(*x*′) *− w*(*y*′).

At the same time, the total number of components equal to zero in *w*(*u*), *w*(*v*) is larger or equal to the number of zero components in *w*(*x*), *w*(*y*), if there are non-zero components in *w*(*x*), *w*(*y*). For, if both *w*_*i*_(*x*′), *w*(*y*′) are different from zero, then either *w*_*i*_(*u*) = *w*_*i*_(*x*′) *− s*_*i*_ will be zero or *w*_*i*_(*v*) = *w*_*i*_(*y*′) *− s*_*i*_, depending on the value of *s*_*i*_ (one zero is added to the total number of zeros). If on the other hand one of *w*_*i*_(*x*′), *w*_*i*_(*y*′) is zero, then *w*_*i*_(*u*) will be zero or *w*_*i*_(*v*), again depending on the value of *s*_*i*_ (no zero is added or omitted).

### 2. “⟹“

#### 2.1 First to the “⟹“-part of the first statement

Let *x, y* be two states with Σ_*i*_ *w*_*i*_(*x*) = Σ_*i*_ *w*_*i*_(*y*). We will show that the observer is indifferent between *x, y*. For this apply *C* a number of times, *n*, to itself until we arrive at two zero vectors *u, v*, i.e. *C*^*n*^(*x, y*) = (*u, v*) with *w*(*u*) = *w*(*v*) = 0. Note that after the re-arrangement step of *C* it is impossible that one of the first components is zero and the other is not, otherwise both vectors would not sum up to the same value. This means, that a zero is added to the total number of zeros as long as one of the first components is non-zero. But the total number of zeros is bounded from above. Thus, after a sufficient number of applications, we get two zero vectors. By (Eudaimonism), the observer is indifferent between *u, v*, i.e. *J* = *I*.

#### 2.2 Now to the “⟹“-part of the second statement

Assume that Σ_*i*_ *w*_*i*_(*x*) *>* Σ_*i*_ *w*_*i*_(*y*). Take the difference between the bigger and the smaller sum and divide it by the number of agents, *n*, yielding 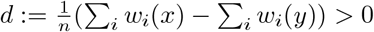

Choose with (Maximum Domain), a new state *v* with *w*_*i*_(*v*) = *w*_*i*_(*x*) *− d*. Observe that Σ Σ_*i*_ *w*_*i*_(*v*) = Σ_*i*_ *w*_*i*_(*y*). By the preceding paragraph we know that *v* is indifferent to *y*, i.e. *vIy*. But by (Unanimity) *v* is strictly preferred to *y*, i.e. *vPy*, since every component in *w*(*x*) is equal or bigger than *w*(*v*) by construction. Therefore, by the compatibility of *I* and *P* we have *xPy*.

### 3. “ ⟸ “

From the preceding we know that a state is preferred to another if the sum of well-being in one state is bigger than in the second; indifferent, if the sums are equal; and less preferred, if the sum of the first state is smaller than that of the second. That is, the judgement respects the ordering of the sums. This implies the “ ⟸ “-part of both statements.

## Data Availability

Code is available online at

https://doi.org/10.5281/zenodo.13336168

## Acknowledgments

I acknowledge the fruitful discussions along the presentations of the worin our joint workshops of the four research groups at the University of Koblenz. Additionally, I would like to thank Prof. Thomas Götz, of the University of Koblenz, and Prof. Tyll Krüger, of the Technical University of Warsaw, and Prof. em. Thomas Kuhn, of the Technical University Chemnitz, for all the fruitful and stimulating discussions we had. I am also indebted to the three anonymous reviewers at this journal for their valuable suggestions to improve the present paper.

## Code Availability

The code used to generated the numerical results can be found in Pestow/Code, 2024.

## Footnotes

2 Given the evidence available at the time, one might have concluded early on that these estimates were exaggerated (Ioannidis, 2020b).

3 Alternatively, in the spirit of the argument above, if a person still believed the early announcement, then the later estimates would demonstrate to that person the existence of widespread bias in the expert community.

4 ”The perceived level of personal threat needs to be increased among those who are complacent, using hard-hitting emotional messaging.” UK Government (2020).

5 ”Emphasize the Worst Case!” Federal Government of Germany (2020); incidentally, COSMO was initiated because “Journalists need timely knowledge about developing audience behaviour and habits to rapidly tailor information sharing and to develop narrative tools that encourage behaviour changes according to evidence from risk communication research.” (Bertsch et al., 2020, coauthored by head of the RKI at that time, the department that was central in the German pandemic management.)

6 The UK health minister at that time wanted to “[…] frighten the pants off everyone with the new strain.” The Cabinet Secretary at that time was of the opinion that in “ramping up messaging - the fear/guilt factor [is] vital” (Lockdown Files, 2023). Scientists involved with the behavioral change effort later questioned the ethicality of the psychological operations employed (Rayner, 2021).

7 Julian Reichelt reported the expectations her publisher had of him as (former) Editor-in-Chief of a widely circulating German tabloid: “Friede Springer had the idea - and made this very clear to me - that Bild should immediately report in support of the German government and the Chancellor in the early stages of the coronavirus crisis. And that was not my idea of journalism.” (Reichelt, 2022)

8 An influential Swiss publisher said in a leaked record that “In all the countries where we are active - and I would be happy if it stays in this circle - we said on my initiative that we want to support the government through our media coverage so that we all get through the crisis well.” (von Matt, 2022)

9 ”[…] messages that provide in-group models for norms (for example, members of your community) may therefore be most effective.”; Effective messaging approaches suggested include “focusing on protecting others (for example, ‘wash your hands to protect your parents and grandparents’), aligning with the recipient’s moral values, appealing to social consensus or scientific norms and/or highlighting social group approval.”; “For instance, a message with compelling social norms might say, ‘the overwhelming majority of people in your community believe that everyone should stay home’.”; “Methods to increase certainty include helping people feel knowledgeable about their new attitude and making them feel that their new attitude is the ‘moral’ one to have.”; “People are also more likely to cooperate when they believe that others are cooperating. […] This suggests that leaders and the media can promote cooperation by making […] [cooperative] behaviours more observable.”; “Leaders can do this, for instance, by being a source of ‘moral elevation’. Visibly displaying prosocial and selfless acts can prompt observers to also act with kindness and generosity themselves.”

10 ”[Psychological] Inoculation follows the biomedical analogy: people are exposed to a severely weakened dose of a persuasive argument, strong enough to trigger the immune system but not so strong as to overwhelm it. A meta-analysis has found inoculation effective in protecting attitudes from persuasion.”; “thus, focusing on worst-case scenarios, even if they are uncertain, may encourage people to make sacrifices for others.”

11 Incidentally, the paper could be useful in the study of sects, like the Peoples Temple, and possibly sudden civilizational collapses as it provides many examples of mind closing mechanisms that were demonstrated to be applicable to larger groups.

12 This axiom is actually a consequence of the previous principle, if one use the identity function as a trivial permutation, *π* = *id*.

13 An alternative argument for evaluating the social state by the sum of well-being or rather the average well-being comes from the contractarian tradition (Harsyani (1955), Sen (1970), Sugden (1979)). We repeat it here for its simplicity. In the thought experiment, as in (Sen (1970)), society is placed behind a “veil-of-ignorance”, which establishes a kind of impersonality. A concept closely related to impartiality. Behind the veil no one knows which position in society they will take or actually have. Since all the characteristics of the individuals behind the veil are hidden, all will take the same decision (as there is nothing to differentiate them) and will therefore agree with each other on one social contract. By Laplaces’ principle of indifference, the probabilities for taking a given position in society are all the same. Thus, under rational choice, in the sense above, individuals will decide on a society where the expected well-being or average well-being is maximized.

14 Furthermore, a rescaling of the sum of well-being does not change the welfare ordering, which means that rescaled sums are valid criteria for the observer, too.

15 The simulations were performed on a 2,3 GHz Intel Core i9 CPU with 16 GB RAM running Python 3.11.1 under MacOS Sonoma 14.5 and take around 15 minutes to generate.

16 The jump discontinuity in the time of peak is probably due to a numerical instability resulting from the discretization of the time axis and the adaptive step size used in integrating the differential equations. We will not rest our analysis on this result.

17 For this and other reasons, the author adheres more to a moral sense tradition of ethics and remains uncommitted about utilitarianism, though very sympathetic to it. We mention this here only to avoid being misidentified as utilitarians.

18 Consider that if the risk of abuse is at least *x*% on any given election, then the risk that this power will be abused at least once after *n* elections is larger than 1 *−* (1 *− x*)^*n*^. This term tends to certainty with increasing *n*, i.e. the risk is certain to be realized if left unaddressed long enough and if *x* is large enough. That the chance *x is* large enough seems to us very plausible in light of the recent Covid-19 pandemic experience.

19 There is one exception which Leibniz noted, namely the empirical fact of the existence of ones consciousness.

20 The above points seem to make the introduction of protective rights against psychological control by the state highly desirable. Otherwise, society is at risk of suffering in terms of damage to democratic legitimacy, polarization, and mal-government (see also (Schippers, 2021, 2022) for other practical implications). The fifth point in particular leads us to emphasize the importance of the practice of *methodic doubt*, especially in science, as this provides a remedy against individual and social errors (e.g. “group think”) and, taken constructively, is a powerful stimulant in the search for scientific truth.

